# Retrospective analysis of SARS-CoV-2 omicron invasion over delta in French regions in 2021-22: a status-based multi-variant model

**DOI:** 10.1101/2022.06.28.22277015

**Authors:** Thomas Haschka, Elisabeta Vergu, Benjamin Roche, Chiara Poletto, Lulla Opatowski

## Abstract

**Background:** SARS-CoV-2 is a rapidly spreading disease affecting human life and the economy on a global scale. The disease has caused so far more then 5.5 million deaths. The omicron outbreak that emerged in Botswana in the south of Africa spread around the globe at further increased rates, and caused unprecedented SARS-CoV-2 infection incidences in several countries. At the start of December 2021 the first omicron cases were reported in France.

**Methods:** In this paper we investigate the contagiousness of this novel variant relatively to the delta variant that was also in circulation in France at that time. Using a dynamic multi-variant model accounting for cross-immunity through a status-based approach, we analyze screening data reported by *Santé Publique France* over 13 metropolitan French regions between 1st of December 2021 and the 30th of January 2022. During the investigated period, the delta variant was replaced by omicron in all metropolitan regions in approximately three weeks. The analysis conducted retrospectively allows us to consider the whole replacement time window and compare regions with different times of omicron introduction and baseline levels of variants’ transmission potential. As large uncertainties regarding cross-immunity among variants persist, uncertainty analyses were carried out to assess its impact on our estimations.

**Results:** Assuming that 80% of the population was immunized against delta, a cross delta/omicron cross-immunity of 25% and omicron generation time was 3.5 days, the relative strength of omicron to delta, expressed as the ratio of their respective reproduction rates,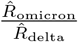, was found to range between 1.51 and 1.86 across regions. Uncertainty analysis on epidemiological parameters led 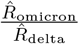 ranging over 1.57-2.13 when averaged over the metropolitan French regions, weighting by population size.

**Conclusions:** Upon introduction, omicron spread rapidly through the French territory and showed a high fitness relative to delta. We documented considerable geographical heterogeneities on the spreading dynamics. The historical reconstruction of variant emergence dynamics provide valuable ground knowledge to face future variant emergence events.

## 1 Background

The SARS-CoV-2 pandemic first emerged in China in December 2019 and subsequently spread all over the world. Despite unprecedented control measures and availability of several vaccines, the virus persisted in populations and evolved into different lineages. These SARS-CoV2 mutations have allowed a classification into different variants that have caused further isolated or overlapping epidemic waves in many countries [1]. In particular, variants with increased transmissibility, increased severity or immune escape were classified as variants of concern (VOC) and named after the letters of the Greek alphabet.

The omicron SARS-CoV-2 VOC, first detected in Botswana in the south of Africa [2] in November 2019, rapidly spread around the world [3, 4]. The mutations observed on this variant are expressing a different form of the Spike protein [5], seemingly causing immune escape [6] and higher infection rates [7, 8, 9, 10].

Omicron was detected for the first time in France at the beginning of December 2021 [11]. Early assessments of its dynamics pointed to a rapid growth, a substantial spreading advantage over the delta variant, the circulating variant at that time in France [12, 13]. Therefore, omicron has been attributed the potential to cause a large-scale epidemic wave [12, 13, 14]. The rate of daily detected cases, indeed, underwent unprecedented growth and over 300 000 detected cases per day were registered in the first half of January for this country consisting of a population of almost 67 million inhabitants [15]. Here, we retrospectively analysed the dynamics of the emergence of the omicron variant and the replacement of the delta one across all French regions. We proposed a multi-variant compartmental model accounting for cross-immunity through a status-based formulation and analysed longitudinal data between December 2021 to January 2022, spanning the entire replacement period. We aimed at quantifying the relative advantage of omicron with respect to delta and its spatial heterogeneity on the replacement dynamics, by accounting for uncertainty in different factors, such as the variants’ generation time.

## 2 Methods

### 2.1 Data Acquisition and preprocessing

PCR-confirmed cases associated with SARS-CoV-2 mutations were obtained from *Santé Publique France* ^1^ for the 13 metropolitan French regions. The data includes the number of tests that underwent the screening for a selection of mutations in their amino acid sequence. Different mutations were monitored for their impact on viral functioning and because they were recognised as indicators of different VOCs. In particular, the E484K mutation is commonly used as an indicator of a beta or gamma variant and the L452R as an indicator of the delta variant. The absence of these two mutations is characteristic of the alpha, omicron or other lineages, e.g. B.1.640. Periodic whole genome sequencing surveys showed that B.1.640 was circulating at a low level in France at the time of omicron introduction to be rapidly replaced by omicron around mid December [12, 13]. In late November 2021, a dedicated surveillance protocol was established in France targeting a set of mutations specific to the omicron variant. The initial set of mutations was soon updated to become in late December: deletion of site 69/70 and/or substitutions K417N and/or S371L-S373P and/or Q493R [16]. The protocol was initially adopted by certain laboratories to become progressively more widespread throughout December, early January.

Available records were used to describe the co-circulation of omicron, delta and beta/gamma. Records with L452R mutation were interpreted as delta variant. E484K mutations were taken as indicator of beta/gamma variant. These were counted in negligible fractions but were kept for completeness. For omicron the two alternative options for data interpretations were subject to different potential biases. The use of the absence of L452R and E484K mutations as a proxy for omicron was biased around the onset of omicron invasion due to the co-circulation with other lineages. Given that omicron became dominant among the samples without L452R mutation at the very beginning of its invasion, as explained above, this biases had likely a limited effect on the whole replacement curve. On the other hand, the use of the omicron-specific set of mutations was likely biased during the period from the end of November and beginning of January when such an indicator was adjusted and gradually adopted throughout the French territory. Given our interest on the entire replacement period between the beginning of December and the end of January, we assumed in the baseline analysis omicron to be described by the absence of L452R and E484K mutations. Still, we considered the alternative indicator in the sensitivity analysis.

Visual inspection of the time series between 1st December 2021 and 31th January 2022 reveals that the invasion of omicron occurs approximately in three weeks in all the regions of metropolitan France. As such we defined for each region a window of opportunity of 20 days for the analysis. The onset of this window is defined by its midpoint, the 10th day, where omicron shall supplant the delta variant in absolute numbers, meaning that the omicron variant exceeds 50% of reported samples at this midpoint. This approach is outlined in figure 1 which represents the data of a typical French region and its selected 20 day window of opportunity (figure 1C). The dataset provided by *Santé Publique France* is smoothed over a 7-day sliding window.

**Figure 1:**
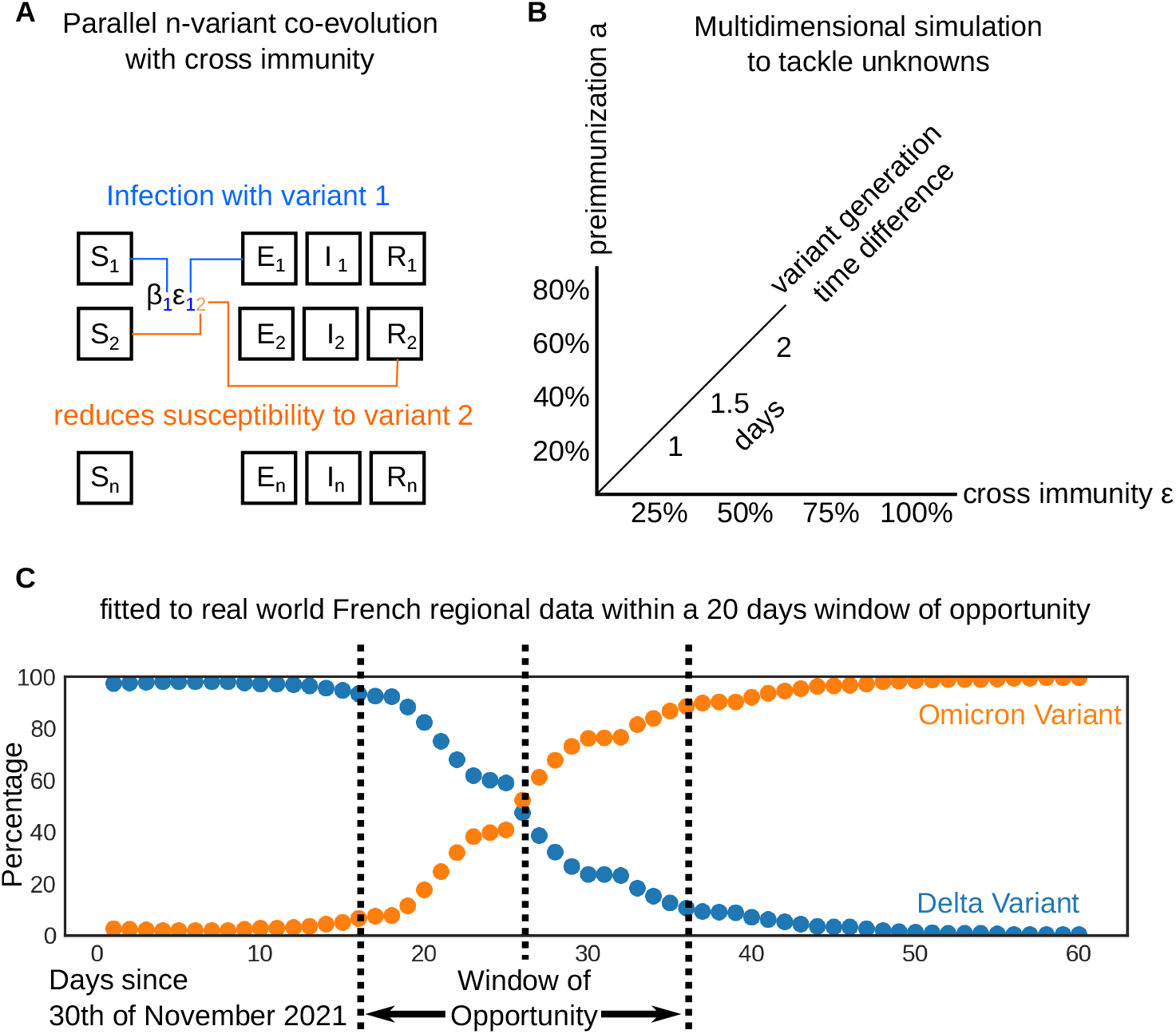
**A**. A basic overview of the model. **B**. The model paramterization possibilities. **C**. As an illustration, percentages of delta and omicron obtained from *Santé Publique France* for the Ile-de-France region with the 20 day window of opportunity around the inflection point as it has been chosen for this modelling study.

### 2.2 Multi-variant transmission model

We modelled the co-circulation of three variants. Inspired by [17], we proposed a status-based multi-variant compartmental model allowing us to evaluate the three variants (delta, omicron and residual beta/gamma) simultaneously, which interact with each other using a cross-immunity term (a schematic view of the model in figure 1A). The model is defined by the ordinary differential equations (1-4), where state variables stand for proportions of different compartments in the population from the viewpoint of each variant *i*:

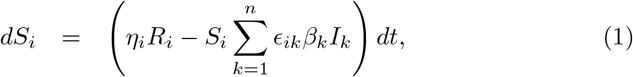

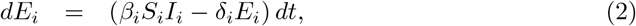

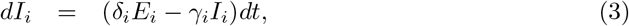

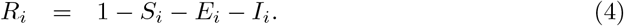

*S*_*i*_ represents the population susceptible, *E*_*i*_ the incubating non infectious population and *I*_*i*_ the population of infectious individuals. Compartment *R*_*i*_ models an immunized population that either underwent infection and recovered from the disease or has been vaccinated. *β*_*i*_ represents the transmission rate, *η*_*i*_ is the immunity waning rate, *δ*_*i*_ the rate at which exposed individuals become infectious, or the inverse of the mean sojourn time in *E* compartment, and *γ*_*i*_ the recovery rate or the inverse of the infectiousness duration. Variant interaction is modelled by a cross-immunity matrix, where element *ϵ*_*ik*_ describes the acquired protection to an acquisition of variant *i* conferred by an infection with variant *k*. For a given reproduction rate at time 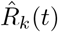, the transmission rate *β*_*k*_ can be obtained from:

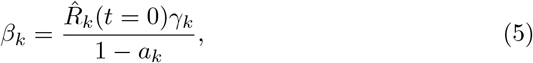

where *a*_*k*_ represents the immunization level in the population against variant *k* at the beginning of the study period.

In the case of the omicron variant, the immunized fraction *a*_omicron_ is obtained by multiplying the fraction of population immune against the delta variant *a*_delta_ with the cross-immunity between omicron and delta:

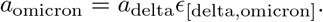

We assume that 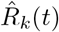 is constant over the investigated period, and we define the relative epidemic fitness of variant *i* to the delta variant as the ratio of reproduction rates: 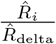

Parameter values were either (i) based on literature values in the case of *δ*_*i*_ and *γ*_*i*_, or (ii) hypothesised for *a*_*k*_, *η*_*i*_ and *ϵ*_*ik*_, with different values tested for robustness purposes for both (i) and (ii) (see section 2.4), or (iii) estimated from data for *β*_*i*_ (related to 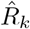, equation (5)). In the baseline scenario, the mean generation time, which is expressed in our model for a variant *i* as 1*/δ*_*i*_ + 1*/γ*_*i*_, was assumed equal to 5 days for delta, 3.5 days for omicron [18] and 8 days for the other variants (beta/gamma) [19, 20]. The corresponding durations (1*/δ*_*i*_ and 1*/γ*_*i*_) in *E*_*i*_ and *I*_*i*_ compartments were assumed equal to (2,3) days, (1.4, 2.1) days and (5,3) days for delta, omicron and beta/gamma variants, respectively.

The model is integrated using a Runge-Kutta-Fehlberg (RKF) algorithm with variable step size [21]. Further we interpolate a third order polynomial on subsequent successions of four obtained data points. This allows us to extract a value on the continuum between the start and the end of the simulation performed with the model described herein. The source code for the model, written in C, has been made available on Github^2^.

### 2.3 Fitting the model to the data and initial conditions

The model was fitted to the reported proportions of sampled variants for each French metropolitan region independently. The fit was performed on a 20 days window starting 10 days prior the inflection point where the omicron variant is present in more than half of the sampled samples.

For all metropolitan French regions, initial conditions were obtained by collecting public estimates of SARS-CoV-2 incidence and reproduction rates, for all variants combined, for the 6th, 13th and 20th of December from *Santé Publique France*^3^. More precisely, the initial conditions were computed based on this data as follows: for each region, *t* = 0 represents the first day of the 20-day study period in our simulations. Depending on the date of the initial point of the study period, we define *q*(*t* = 0) and 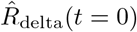 to be the linear interpolations of these collected estimates for each region. If the first day of the 20 days of the simulated window happens before the 6th of December or after the 20th of December, no interpolation is performed and the values from the respective day are taken. Visual inspection confirms that 10 days prior the inflection point, at the beginning of our window of opportunity, omicron cases were still very rare. As such, the interpolated reproduction rate 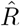 was entirely attributed to the delta variant and stayed constant through the study period.

For each region-associated time window, we used the obtained initial incidence rate *q*(*t* = 0) to set the initial conditions of compartments *E*_*i*_(*t* = 0) and *I*_*i*_(*t* = 0) as follows:

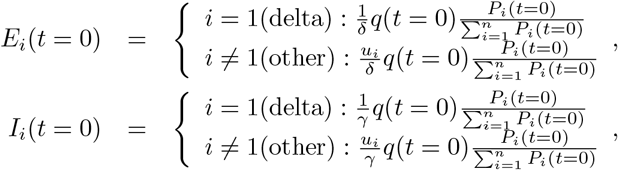

where *P*_*i*_ is the number of positive samples of variant *i* found in the data at the onset date of our study period. *u*_*i*_ is a fit parameter which defines the initial proportion, for each variant *i* except delta variant.

Curve fitting was achieved using a standard gradient descent procedure. Parameters related to VOCs other than the delta variant, which was kept static, were optimized by minimizing the following loss function independently for each region:

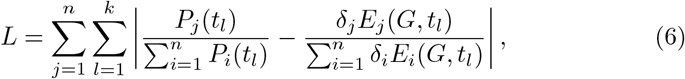

for *n* variants and *k* sampled moments in time. Here *P*_*i*_ represents the observed data, as defined above, and *δ*_*j*_*E*_*j*_(*G, t*_*l*_) the simulated incidence of new infectious individuals at time point *t*_*l*_ as described by equation (2), where 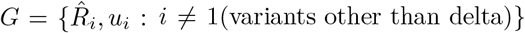. Parameters minimizing the loss function *L* (6) were estimated. Mean estimates as well as standard variations across regions for the omicron reproduction ratio were calculated. We also computed estimates weighted according to the regions population size:

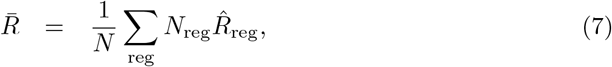

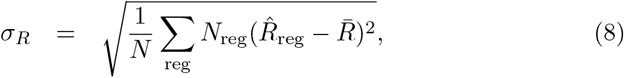

with *N* representing the total population of metropolitan France and *N*_reg_ the population of a single region *reg*. 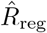 is the best result yielded by the gradient descent for 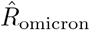 obtained for the corresponding region.

As outlined in figure 1B, independent fits were performed in conjunction with parameter sweeps to overcome uncertainties as further detailed in section 2.4.

### 2.4 Sensitivity analysis

As uncertainty still exists regarding some model parameters such as the generation time and few information is available on others such as variant-specific immunity in the population, we performed a sensitivity/uncertainty analysis to investigate the impact of the different model parameters on our estimates and subsequent variant dynamics.

More precisely, we varied: (i) the generation time of the omicron variant setting it to equal to 3, 3.5 and 4 days as shown in table 1 which summarizes tested values, (ii) the acquired immunity in the population *a* against variants prior to omicron obtained either by infection or vaccination, with *a* ∈ (0.2, 0.4, 0.6, 0.8), and (iii) the cross-immunity *ϵ* that this immunity confers to the omicron variant, with *ϵ* ∈ (0.25, 0.5, 0.75, 1.0).

**Table 1:** Values used in the sensitivity analysis for mean generation time and duration in *E* and *I* compartments for each variant, from [18, 19, 20]. All values are given in days

| Description | Delta | Omicron |
| --- | --- | --- |
| **mean generation time** | 5 | 3 |
| **mean duration in $E$ $1/\delta$** | 2 | 1.2 |
| **mean infectiousness duration $1/\gamma$** | 3 | 1.8 |
| **mean generation time** | 5 | 3.5 |
| **mean duration in $E$ $1/\delta$** | 2 | 1.4 |
| **mean infectiousness duration $1/\gamma$** | 3 | 2.1 |
| **mean generation time** | 5 | 4 |
| **mean duration in $E$ $1/\delta$** | 2 | 1.6 |
| **mean infectiousness duration $1/\gamma$** | 3 | 2.4 |
| **mean immunity duration $1/\eta$** | 1000 | 1000 |

First, the impact of varying model parameters in terms of model fitting was explored for all parameters (i)-(iii).

Second, the impact of the uncertainty in model parameters on replacement dynamics was investigated. When a new variant replaces the established variant, we can numerically quantify the relative fitness by means of Δ*t*, i.e. the time it takes for a new variant to rise from 10% to 50% of positively tested cases in a population. This idea is illustrated in figure 4A, for both a less fitter new variant (orange) characterised by Δ*t*_1_ and a stronger new variant (blue) characterised by Δ*t*_2_, respectively. To get a better understanding of the relationship between our fitness estimates and parameters *a* (ii) and *ϵ* (iii), we analysed in the model variations of:

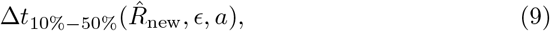

as a function of the reproduction rate 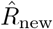 of the new variant, the cross-immunity between invading and established variant *ϵ* and the immunity *a* against the established variant. For these simulations, we fixed the reproduction rate 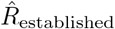 of the established variant to 1.1. and evaluated our model on a grid varying 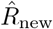, *ϵ* and *a* to evaluate resulting Δ*t*. 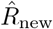 was varied between 1.2 and 2.2 in 31 increments of 0.34, whereas *ϵ* and *a* were given each four different values as detailed above, yielding a total of 512 simulation scenarios.

## 3 Results

### 3.1 Regional fits and relative fitness of omicron against delta in metropolitan France

Assuming that 80% of the population was immunized against delta either by natural infection or vaccination, *a* = 0.8, that one fourth of this immune population procured an immunity against omicron, *ϵ*_[delta,omicron]_ = 0.25, and that generation time was of 5 days for the delta and 3.5 days for omicron variant respectively, the relative fitness of the omicron variant over the delta variant 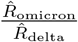 is estimated to be 1.724 (weighted mean over all regions).

For this specific scenario, region specific assumed values for 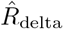 ans estimated values for 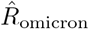 are displayed in table S1, and replacement curves and associated fits for all regions are reported in figure 2. The general trend is globally well reproduced by the model in all regions. The lowest fitness estimate was obtained for Corsica, with 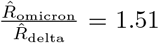, while the highest for Nouvelle-Aquitaine, with 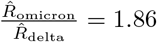. Distribution of fitness values is summarised in figure 3. These heterogeneous values were not correlated to population density (figure S7).

**Figure 2:**
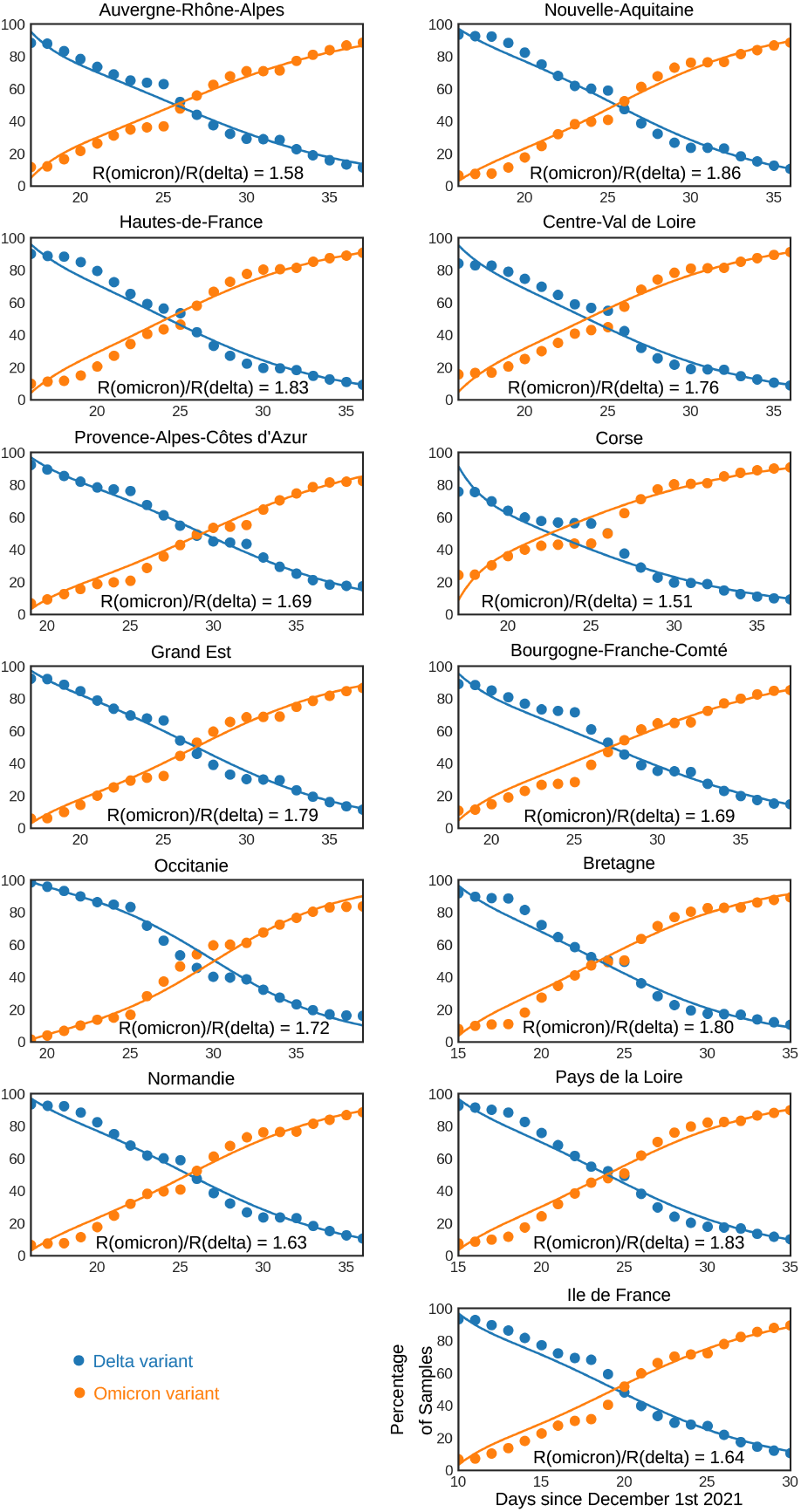
Omicron and delta SARS-CoV-2 variant proportions among positive samples in the regions respective window of opportunity, 10 days before and after the omicron variant exceeds 50%. Dots are representing proportions reported from sampled data published by *Santé Publique France*. Lines represent simulated data with estimated parameter values, here with a delta-omicron cross-immunity of 25% and an initial population that is 80% immunized against the delta variant. The generation time assumed here is 5 days for the delta variant and 3.5 days for the omicron variant.

**Figure 3:**
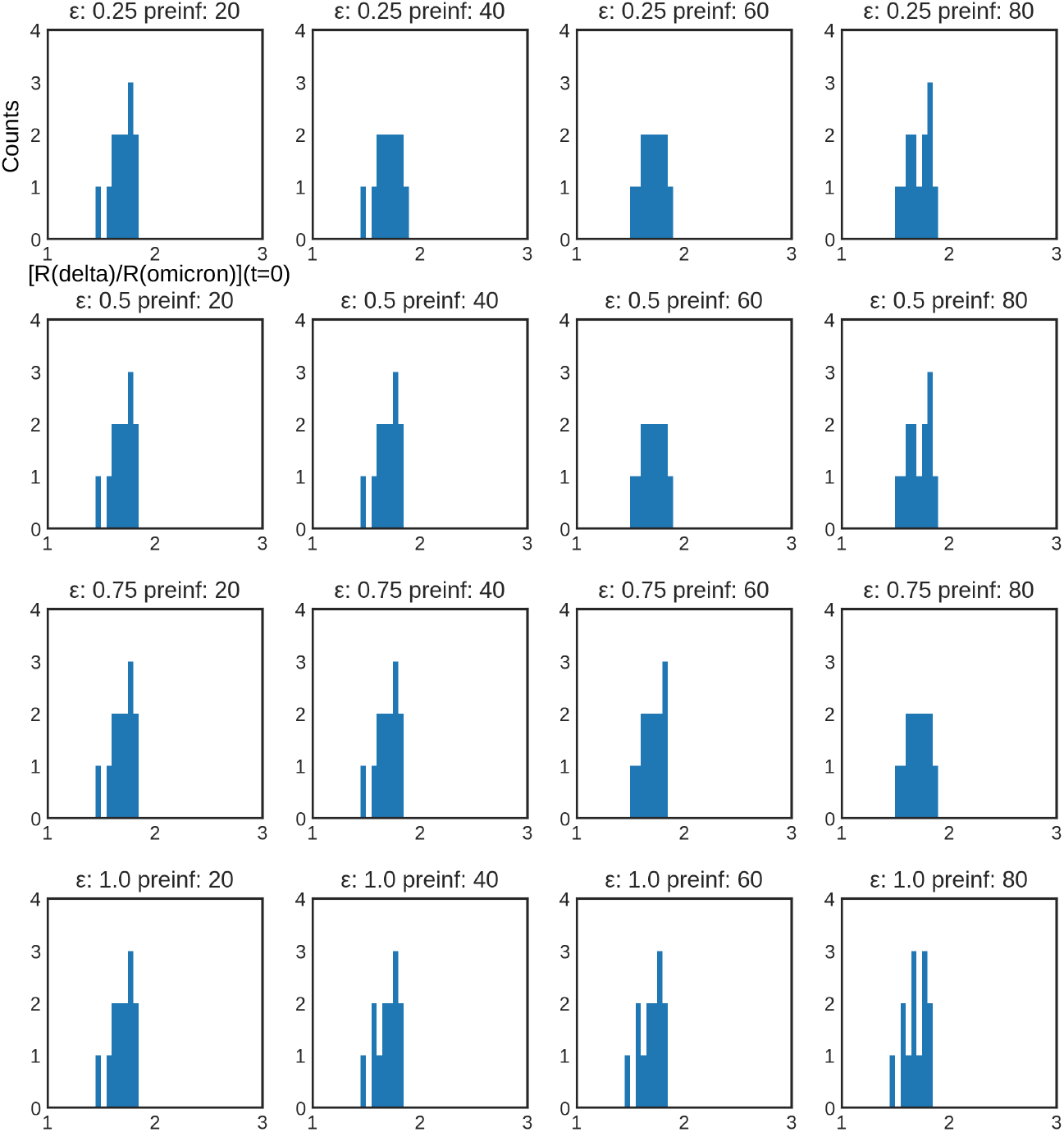
Distribution of 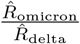 by regions shown by a 40 bins histogram between the values 1 and 3, when varying the cross-immunity *ϵ* and preimmunization level *a* (preinf). The generation time was 5 days for the delta variant and 3.5 days for the omicron variant.

### 3.2 Sensitivity to uncertain model parameters

As a result of the robustness analysis on fitting, when the generation time of omicron is sampled at 3, 3.5 and 4 days, we can remark that the fitness of the omicron variant increases as the generation time approaches the generation time of the delta variant. The resulting shift in relative fitness can be observed in comparing figures 3 and S1 to S5 (S3-S5 when using data with the alternative definition of omicron) found in the supporting information.

During our parameter sweep, the minimal relative fitness 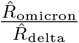 of 1.38 is observed for Corsica as the generation time for omicron is set to 3 days. The Nouvelle-Aquitaine region exhibits the maximum value of 1.99 as the generation time for omicron is 4 days. Assuming a generation time of 5 days for delta and 3 days for the omicron variant we find that the average relative fitness 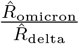, weighted by regional population size, ranged from 1.58 to 1.61. Increasing the generation time of omicron from 3 to 4, the relative fitness is increased to 1.81-1.84 in average. Estimates according to generation time assumptions are detailed in table 2.

**Table 2:** Resulting estimates for a range of scenarios varying delta-to-variant cross-immunity (*ϵ*), preimmunized populations proportion (*a*) and generation time (GT). The table displays 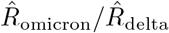 values (weighted mean across regions). Further minimum and maximum values for variance across regions as well as means of the loss function (6), both weighted by population size, are outlined at the bottom of each generation-time associated scenario.

| | $a = 0.2$ | $a = 0.4$ | $a = 0.6$ | $a = 0.8$ |
| --- | --- | --- | --- | --- |
|  | **GT(delta 5, omicron 3) days** |  |  |  |
| $\epsilon = 0.25$ | 1.589 | 1.591 | 1.595 | 1.607 |
| $\epsilon = 0.5$ | 1.592 | 1.565 | 1.594 | 1.608 |
| $\epsilon = 0.75$ | 1.590 | 1.590 | 1.593 | 1.603 |
| $\epsilon = 1.0$ | 1.588 | 1.587 | 1.585 | 1.580 |
| Variance | $\sigma^2 = [0.006 - 0.022]$ | | | |
| Loss | $L = [1.26 - 1.85]$ | | | |
|  | **GT(delta 5, omicron 3.5) days** |  |  |  |
| $\epsilon = 0.25$ | 1.705 | 1.707 | 1.711 | 1.724 |
| $\epsilon = 0.5$ | 1.704 | 1.706 | 1.711 | 1.725 |
| $\epsilon = 0.75$ | 1.705 | 1.705 | 1.707 | 1.720 |
| $\epsilon = 1.0$ | 1.704 | 1.704 | 1.702 | 1.696 |
| Variance | $\sigma^2 = [0.008 - 0.009]$ | | | |
| Loss | $L = [1.39 - 1.42]$ | | | |
|  | **GT(delta 5, omicron 4) days** |  |  |  |
| $\epsilon = 0.25$ | 1.823 | 1.824 | 1.827 | 1.838 |
| $\epsilon = 0.5$ | 1.823 | 1.823 | 1.826 | 1.838 |
| $\epsilon = 0.75$ | 1.823 | 1.823 | 1.824 | 1.835 |
| $\epsilon = 1.0$ | 1.823 | 1.822 | 1.820 | 1.814 |
| Variance | $\sigma^2 = [0.012 - 0.013]$ | | | |
| Loss | $L = [1.48 - 1.54]$ | | | |

Estimates of omicron relative fitness do not seem to depend on the assumptions on cross-immunity conferred by previous immunity to delta in preventing omicron acquisition (table 2 and figure 3). Varying the preimmunization levels of the population at the onset of the omicron invasion does not affect relative fitness values obtained from our model either. When varying these parameters, resulting fits and errors are similar to the ones shown in figure 2. The detailed outputs at region level have been made available on the Github repository^4^.

Note that, despite no effect of cross-immunity on omicron relative fitness was observed here, the corresponding transmission rate of the model *β*, as outlined in equation (5), varies with variations of preimmunization *a* against delta and cross immunity *ϵ* by the predetermined factor 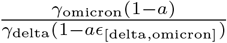, even if almost constant relative fitness rates 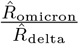 are reported in table 2.

Figure 4B outlines the expected values of Δ*t* as a function of the variant parameters as defined in equation (9). Simulations show that, at relatively high fitness levels, as those found in real omicron and delta variant data 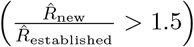, immunity and cross-immunity play a negligible role in the time needed, Δ*t*, to reach the replacement of an established variant by a new fitter variant. In simulations, as 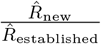 rises, variations in preimmunization *a* and cross-immunity *ϵ* values become irrelevant and do not influence the time needed for a new variant to supplant an established one.

**Figure 4:**
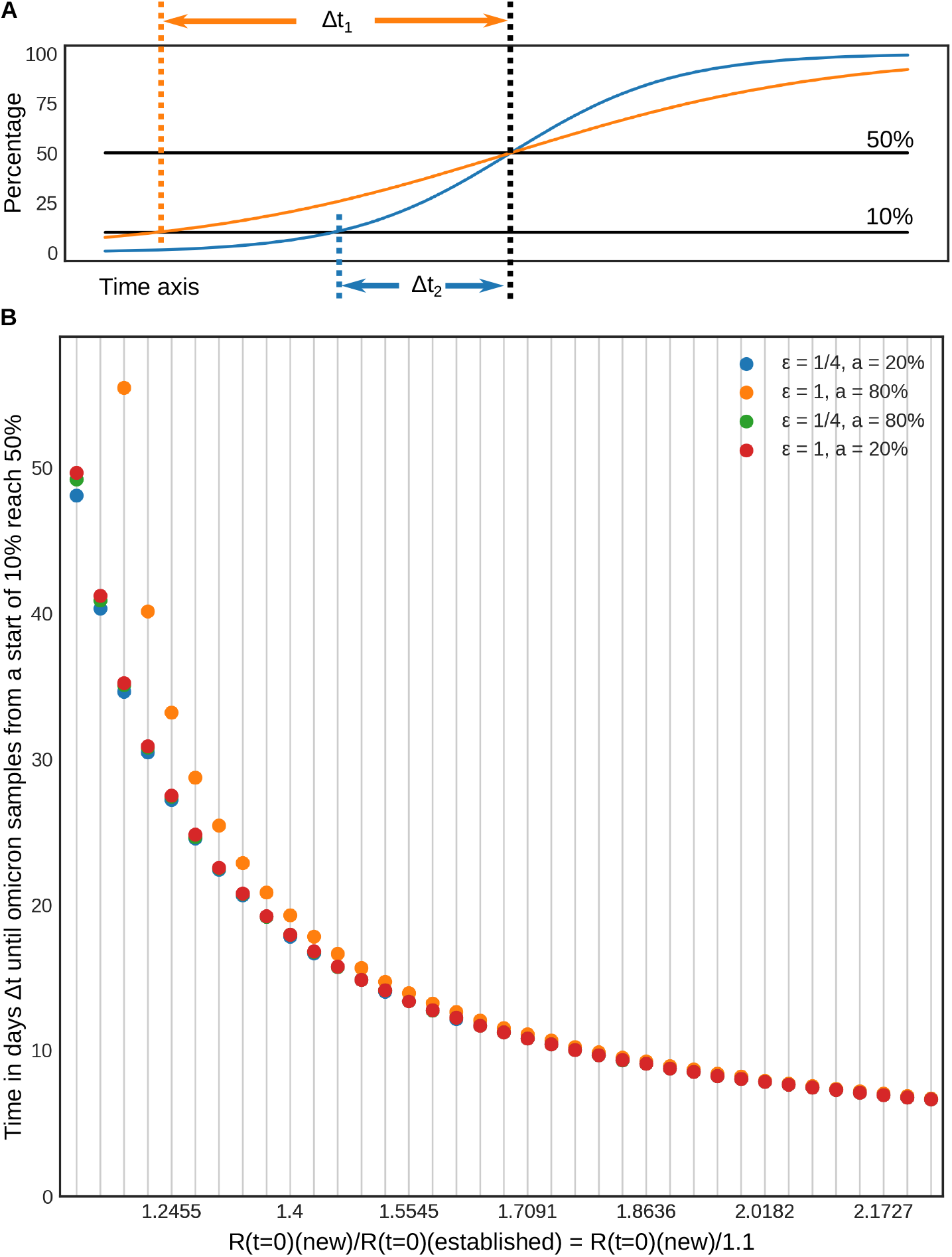
**A**. Illustration for Δ*t*_10%*−*50%_ (defined in equation (9)) taking two examples: a less fitter new variant (orange, Δ*t*_1_) and stronger new variant (blue, Δ*t*_2_) curves, respectively. **B**. Δ*t*_10%*−*50%_ for different relative fitness 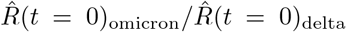, preimmunization (*a*) and cross-immunity (*ϵ*) values. We see that in the regime of 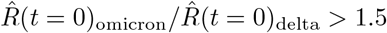, preimmunization and cross-immunity play a minor role in the steepness of variant replacement. On x-axis labels, “new” ans “established” stand for omicron and delta, respectively.

## 4 Discussion

In this article we present a mechanistic model to simulate a multivariant epidemic of SARS-CoV-2. The model was used to analyse SARS-CoV-2 variant data reported by *Santé Publique France* between 1st of December 2021 and 30th of January 2022 on the proportions of different mutations occurring in a specially sampled subset of PCR tests at the French regional level. We detail how this model can be applied in order to evaluate the fitness of an emerging variant, relatively to established SARS-CoV-2 variants. Knowing these values when a new variant is introduced into a population is important for decision-makers in order to evaluate risks caused by the epidemic and carefully plan future stress exhorted on public health systems, economy and other affected areas.

Using the modeling framework outlined herein, we quantify the transmissibility advantage of the omicron variant as outlined in table 2 and figures 2 and 3. The relative fitness between the omicron and delta variant is expressed by the fraction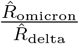 and was found to range between 1.51 and 1.86 across regions when assuming that 80% of the population was immunized against delta, a cross delta_omicron cross-immunity of 25% and omicron generation time was 3.5 days.

Figure 3 displays the increased transmissibility of the omicron variant compared to the delta variant as a distribution across all French regions. Using various parameter sweeps, we showed that this result was very robust and is almost not influenced by different hypotheses about the delta-omicron cross-immunity or preimmunity level to the delta variant at the beginning of December (table 2). Across all scenarios, our weighted average estimates of the increased transmissibility found for the omicron variant compared to the delta variant ranged from 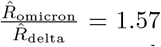 to 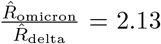 over metropolitan French regions. It is worth mentioning that these results were obtained based on the absence of the L452R mutation only to identify omicron. Results for different variant indicators to describe omicron are highlighted in table S2 and figures S3-S5 in the supporting information.

Two studies provided early assessment of omicron initial spread in France [12, 13]. The study in [13] quantified the doubling time of omicron at the national level and for the Île de France and Centre-Val de Loire regions - the only two regions reporting substantial spread in the community at the time of the study - reporting values ranging between 1.8 ad 2.5 days. The additive advantage in transmission was estimated at 105% by [12] from nationwide data. Our analysis considers the whole period of omicron replacement in all 13 regions in metropolitan France. Results confirm the rapid spread of omicron and provide estimates compatible with [12]. We highlight a variation in omicron fitness by region with a 20% deviation between the minimum and maximum transmission advantage relative to the weighted average across regions.

Our analysis is also comparable with previous studies made outside of France. Similar estimates of main parameters were indeed provided for Great Britain [18]. The British study finds an implied transmission advantage of omicron in the range of 160%-210%. Prior studies from South Africa data find values either slightly above ours 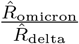 [7] ranging from 1.8 to 3.2, with a mean variation over the month of November 2021 to be between 2.3 and 2.6, or in a second study [8] below our results with values for 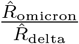 lying between 0.75 and 2.0. Interestingly, a study using Danish data [9] estimated much higher values 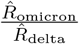 3.19 with a 95% confidence interval ranging from 2.82 to 3.61. Differences in the estimates may depend on different factors, including the surveillance protocols, the precise definition of the transmission advantage, and the modelling approach used for the data analysis.

The preimmunization levels of the population as well as different hypotheses about delta-omicron cross-immunity have almost no effect on the relative fitness estimates (tables 2 and S2), possibly due also to identifiablity issues. Our simulations supported this result, showing that variations in *ϵ*_[delta,omicron]_ and *a* are not expected to yield significant deviations in the dynamics of variant replacement when 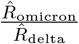 is high enough (*>* 1.5) (figure 4).

The relative fitness 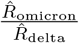 is however sensitive to hypotheses regarding differences in generation time between the delta and omicron variants. We investigated three values (3, 3.5 and 4 days), for the generation time duration of the omicron variant, while keeping the delta variant at a constant generation time of 5 days. Although estimates distributions across regions were not changed, a shift towards higher values of 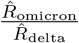 was observed as the generation time for the omicron was variant increased. Results for comparing different generation times are outlined in table 2 and in figure 3 as well as in figures S1-S5 in the supporting information. This result is coherent with a previously published analysis of omicron invasion in Great Britain [18].

During 2021, prior to the omicron wave, several studies have hypothesized different scenarios for the winter period 2021/2022. Sah et al. [22] have built several models to predict the situation in the USA. These authors state that immune escape has to be coupled with increased transmission rates for a variant to be successful. Dyson et al. [23] also explored, through various models, the dynamics of several hypothetical variants. The authors of [23] show that immune escape can slowly develop future waves that might not be easily predictable and can hit the population at later stages of the pandemic. Compared to our study, [22, 23] make long term predictions while we focus on the short period of quick replacement by omicron of the delta variant which lasted, as outlined, only three weeks.

As the analyzed data is not derived from whole genome sequencing but rather from the identification of specific mutations of the virus genome, the data published by *Santé Publique France* that tracks omicron strains allowed for different interpretations of the dataset. Here, we adopted the L452R mutation as an indicator for the delta variant, in presence of the mutation, and the omicron variant, in its absence. An alternative approach consists in combining the mutation L452R taken as a proxy for the delta variant with the set of omicron-specific mutations - deletion of site 69/70 and/or substitutions K417N and/or S371L-S373P and/or Q493R - as an indicator for omicron. The use of the two different indicators yields two distinct time series for each studied regions. Using only L452R both as an indicator for delta, in the presence of the mutation, as well as for omicron, in the absence of the mutation, yields less fluctuations in the time series, as outlined by the comparison of figures 2 and S6. Using only this single mutation may in principle overestimate the of the omicron proportion at the onset of the omicron invasion, as a small non-delta fraction was continuously present in the dataset. However, the bias is likely to be limited in that full genome sequencing data showed that omicron become rapidly dominant among other variants without the L452R mutation during the first weeks of December [12, 13]. The time series based on multi-mutation definition were also analysed and relative fitness values 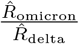 were evaluated. A larger omicron fitness was estimated as we analysed this second time series. This could be explained by the progressive mounting of this surveillance protocol concomitant with the omicron invasion, which may have biased the observation of the omicron dynamics. The different results for the two representations are outlined in the tables 2 and S2. However, both definitions led to estimates that were consistent with those reported by various studies around the globe [7, 9, 8, 18].

There is no significant correlation between population density and effective omicron relative fitness as outlined by figure S7 in the supporting information.

## 5 Conclusion

We estimated that as omicron replaced delta in France during winter 2021/2022, the relative fitness of the omicron variant compared to the delta variant, 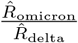, ranges from 1.57 to 2.13. We propose here a multi-variant framework that enables short-term analysis of the epidemiological characteristics of an emerging variant using epidemiological data. The model presented here could be easily applied to other dynamic systems describing the evolution of epidemic diseases evolving into different variants.

## Data Availability

The epidemiological model used to perform the analyses herein has been made available on github
Input data concerning regional variant sampling that has been processed herein is provided by Sante Publique France and is publicly available.
Raw regional results concerning the replacement of the delta variant by the omicron variant are made available at github.

https://github.com/haschka/SIER_multivariant_epidemic

https://www.data.gouv.fr/fr/datasets/donnees-de-laboratoires-pour-le-depistage-indicateurs-sur-les-mutations/

https://github.com/haschka/French-Regional-Omicron-Invasion

## Availability of data and materials

The epidemiological model used to perform the analyses herein is available at https://github.com/haschka/SIER_multivariant_epidemic.

Raw regional results are made available at https://github.com/haschka/French-Regional-Omicron-Invasion.

Input data concerning regional variant sampling that has been processed herein is provided by *Santé Publique France* and available at: https://www.data.gouv.fr/fr/datasets/donnees-de-laboratoires-pour-le-depistage-indicateurs-sur-les-mutations/

## Authors’ contributions

T.H., C.P., B.R, E.V and L.O conceptualized the model and designed the analysis. T.H. implemented the model and estimation algorithm in C. C.P., B.R, E.V and L.O parametrized the model for the simulations. T.H. extracted the data and performed the data analyses. T.H, C.P., B.R, E.V and L.O wrote the manuscript.

## Additional Files

### Additional file 1 — Additional results of sensitivity analysis

This file contains additional tables and figures as follows: regional values of assumed and estimated reproduction ratios for a specific scenario (table S1); results of sensitivity analysis for an alternative interpretation of data (table S2); fitness distribution across regions for different values of the generation interval for omicron with the first interpretation of data (figures S1 ans S2) or the alternative one (figures S3-S5); fits results for all regions with the alternative interpretation of data (figure S6); correlation between population density and estimated relative fitness (figure S7).

## Footnotes

1 https://www.data.gouv.fr/fr/datasets/donnees-de-laboratoires-pour-le-depistage-indicateurs-sur-les-mutations/

2 https://github.com/haschka/SIER_multivariant_epidemic/

3 https://www.santepubliquefrance.fr/dossiers/coronavirus-covid-19/coronavirus-chiffres-cles-et-evolution-de-la-covid-19-en-france-et-dans-le-monde

4 https://github.com/haschka/French-Regional-Omicron-Invasion

