## Supplementary material for "Retrospective analysis of SARS-CoV-2 omicron invasion over delta in French regions in 2021-22: a status-based multi-variant model": Supplimental Information

| Region | Onset Date | $\hat{R}_{\text{delta}}$ | $\hat{R}_{\text{omicron}}$ | Error[††] |
|---|---|---|---|---|
| Auvergne-Rhône-Alpes | 17 | 1.17 | 1.85 | 1.28 |
| Bourgogne-Franche-Comté | 18 | 1.00 | 1.69 | 1.51 |
| Bretagne | 15 | 1.09 | 1.96 | 1.49 |
| Corse | 17 | 1.24 | 1.88 | 2.00 |
| Centre-Val de Loire | 16 | 1.06 | 1.86 | 1.80 |
| Grand Est | 17 | 1.07 | 1.92 | 1.22 |
| Hauts-de-France | 16 | 1.00 | 1.83 | 1.59 |
| Ile-de-France | 10 | 1.27 | 2.08 | 1.63 |
| Nouvelle-Aquitaine | 16 | 1.01 | 1.89 | 1.25 |
| Normandie | 14 | 1.16 | 1.89 | 1.99 |
| Occitanie | 18 | 1.09 | 1.87 | 0.78 |
| Provence-Alpes-Côte d'Azur | 19 | 1.10 | 1.86 | 1.22 |
| Bretagne | 15 | 1.04 | 1.91 | 1.44 |

Table S1: Initial conditions for the 13 regions and corresponding estimates for omicron variant reproduction ratio. The onset date is the day of December 2021 which represents the first day of our 20 day evaluation window for each region.
$\hat{R}_{\text{delta}}$ represents, for each region, the assumed reproduction ratio at onset for Delta variant. Regional estimates of reproduction rates $\hat{R}_{\text{omicron}}$ in the French metropolitan regions are obtained under the assumption of the following parameter values: $\epsilon = 25\%$ for the cross-immunity between the omicron and delta variant, $a = 80\%$ as the population percentage being immune to the delta variant and generation times of 5 days for delta and 3.5 days for omicron respectively. The definition of omicron is based on the **absence of L452R mutation**.
Tables outlining all experiences performed under various assumptions can be found on github:
`https://github.com/haschka/French-Regional-Omicron-Invasion`
[††] Error represents the remaining loss after minimization by gradient descent as defined by equation (6) in the original article.

| | $a = 0.2$ | $a = 0.4$ | $a = 0.6$ | $a = 0.8$ |
|---|---|---|---|---|
| | **GT(delta 5, omicron 3) days** | | | |
| $\epsilon = 0.25$ | 1.807 | 1.809 | 1.812 | 1.823 |
| $\epsilon = 0.5$ | 1.808 | 1.809 | 1.811 | 1.819 |
| $\epsilon = 0.75$ | 1.809 | 1.807 | 1.809 | 1.815 |
| $\epsilon = 1.0$ | 1.809 | 1.806 | 1.804 | 1.797 |
| Variance | $\sigma^2 = [0.079 - 0.085]$ | | | |
| Loss | $L = [1.75 - 1.78]$ | | | |
| | **GT(delta 5, omicron 3.5) days** | | | |
| $\epsilon = 0.25$ | 1.966 | 1.972 | 1.972 | 1.965 |
| $\epsilon = 0.5$ | 1.983 | 1.972 | 1.956 | 1.967 |
| $\epsilon = 0.75$ | 1.970 | 1.952 | 1.973 | 1.965 |
| $\epsilon = 1.0$ | 1.985 | 1.957 | 1.964 | 1.973 |
| Variance | $\sigma^2 = [0.056 - 0.112]$ | | | |
| Loss | $L = [1.72 - 1.80]$ | | | |
| | **GT(delta 5, omicron 4) days** | | | |
| $\epsilon = 0.25$ | 2.114 | 2.107 | 2.114 | 2.125 |
| $\epsilon = 0.5$ | 2.114 | 2.116 | 2.116 | 2.127 |
| $\epsilon = 0.75$ | 2.114 | 2.110 | 2.124 | 2.127 |
| $\epsilon = 1.0$ | 2.108 | 2.107 | 2.108 | 2.098 |
| Variance | $\sigma^2 = [0.068 - 0.095]$ | | | |
| Loss | $L = [1.74 - 1.77]$ | | | |

Table S2: Resulting estimates for a range of scenarios varying delta-to-variant cross immunity ($a$), preimmunized populations proportion and generation times (GT). The table displays $\hat{R}_{\text{omicron}}/\hat{R}_{\text{delta}}$ mean weighted values across regions. Further minumum and maximum values for variance across regions as well as means of the loss function, both weighted by population size, are outlined at the bottom of each generation-time associated scenario. The alternative definition of omicron is based on the following mutations: **deletion of site 69/70 and/or substitution K417N and/or S371L-S373P and/or Q493R**.

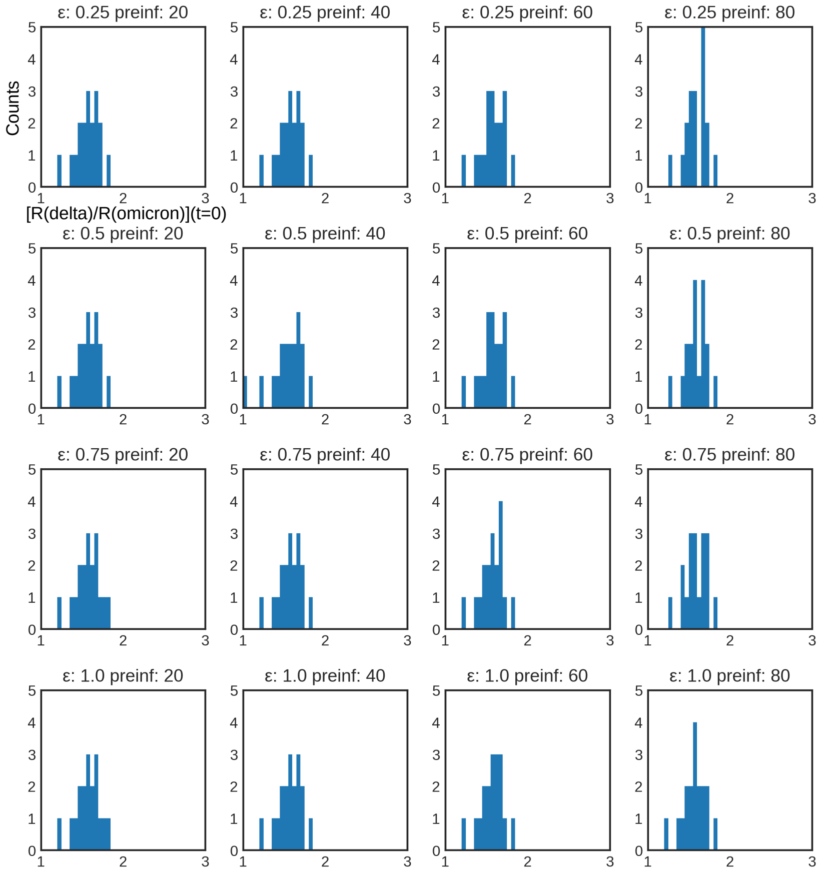


Figure S1: Fitness distribution across regions when assuming omicron generation time to be 3 days on average. The graphic shows the distribution of $\frac{\hat{R}_{\text{omicron}}}{\hat{R}_{\text{delta}}}$ by regions shown by a 40 bins histogram between the values 1 and 3 according to generation times of 5 days for the delta variant and 3 days for the omicron variant. The definition of omicron is based on the **absence of L452R mutation**.

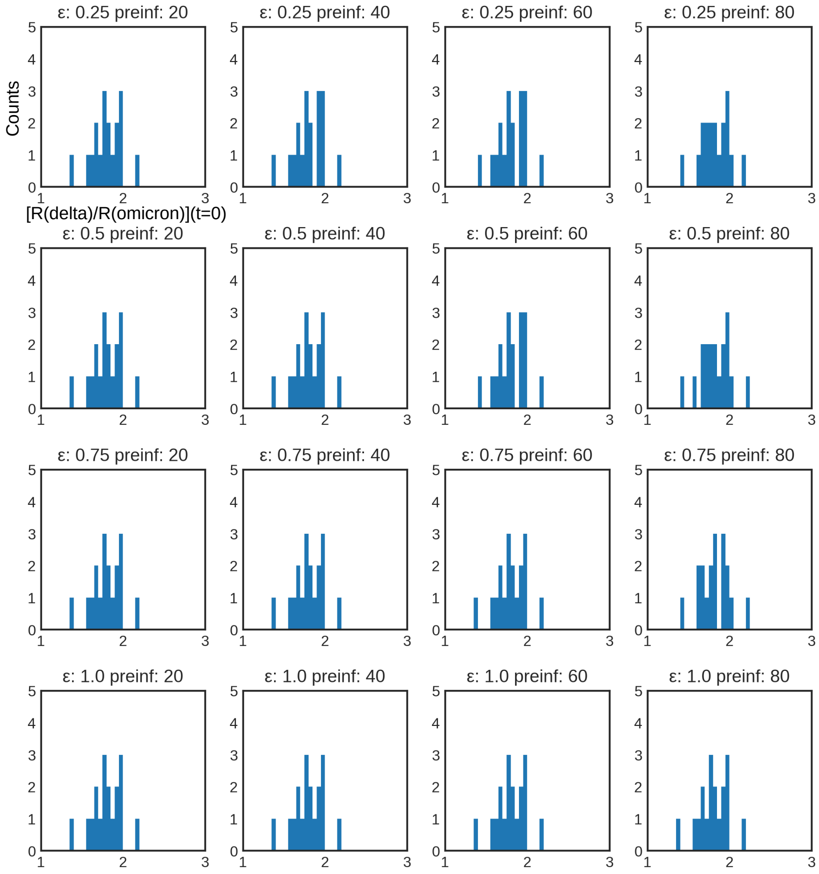


Figure S2: Fitness distribution across regions when assuming an omicron generation time to be 4 days on average. The distribution of $\frac{\hat{R}_{\text{omicron}}}{\hat{R}_{\text{delta}}}$ by regions shown by a 40 bins histogram between the values 1 and 3 according to generation times of 5 days for the delta variant and 4 days for the omicron variant. The definition of omicron is based on the **absence of L452R mutation**.

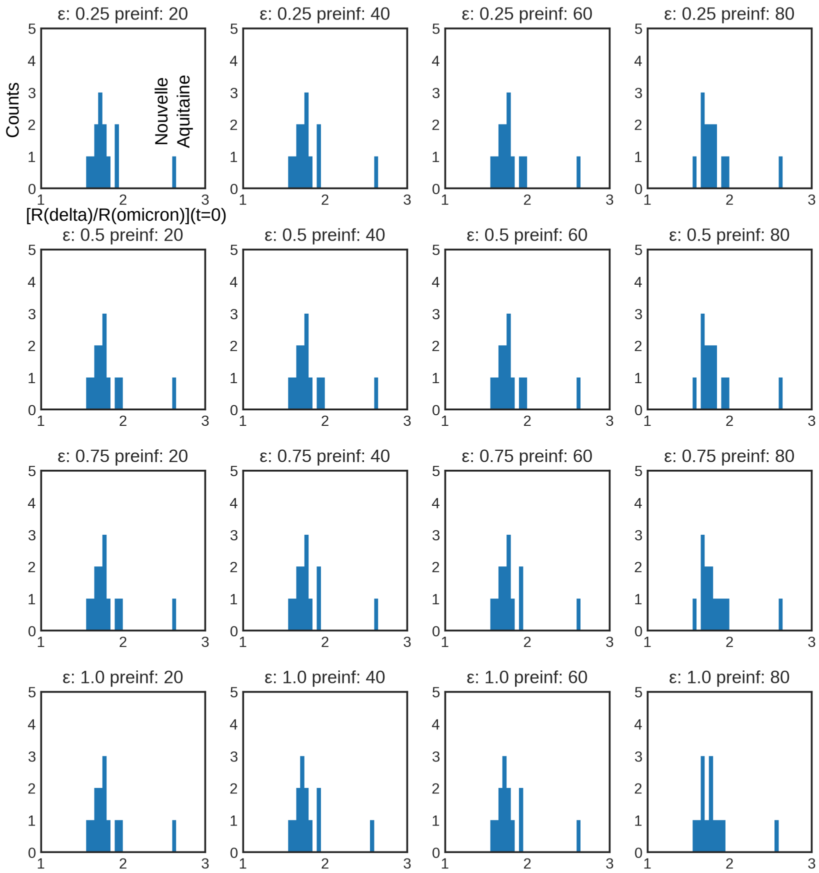


Figure S3: Fitness distribution across regions assuming an omicron generation time of 3 days on average. The distribution of $\frac{\hat{R}_{\text{omicron}}}{\hat{R}_{\text{delta}}}$ by regions shown by a 40 bins histogram between the values 1 and 3 according to generation times of 5 days for the delta variant and 3 days for the omicron variant. The alternative definition of omicron is based on the following mutations: **deletion of site 69/70 and/or substitution K417N and/or S371L-S373P and/or Q493R**.

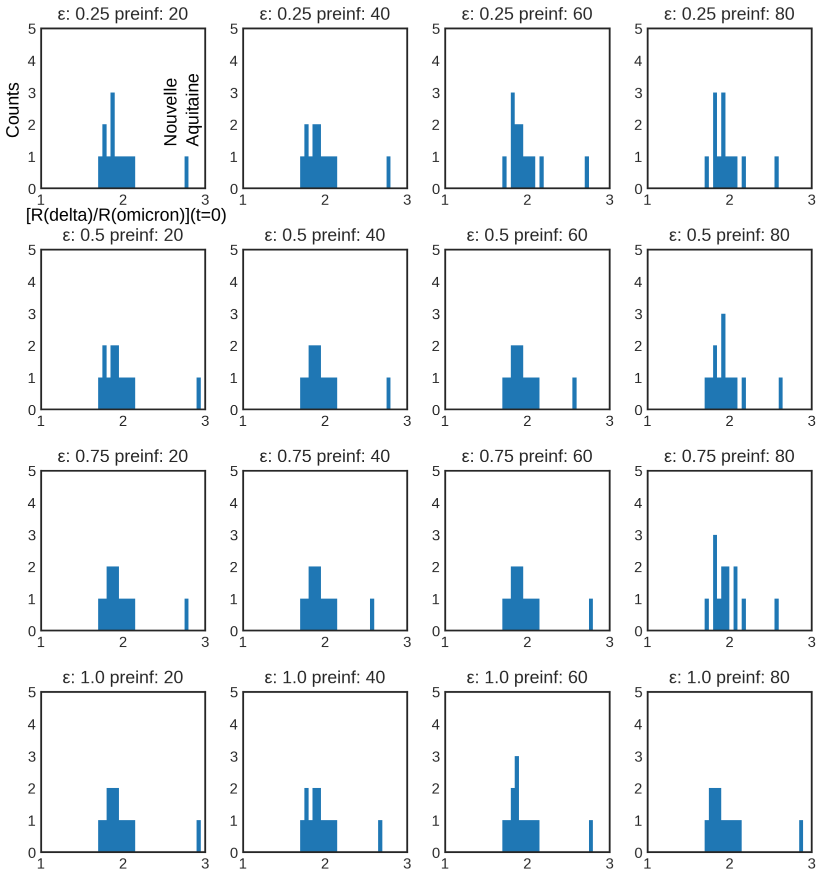


Figure S4: Fitness distribution across regions assuming an omicron generation time of 3.5 days on average. The distribution of $\frac{\hat{R}_{\text{omicron}}}{\hat{R}_{\text{delta}}}$ by regions shown by a 40 bins histogram between the values 1 and 3 according to generation times of 5 days for the delta variant and 3.5 days for the omicron variant. The alternative definition of omicron is based on the following mutations: **deletion of site 69/70 and/or substitution K417N and/or S371L-S373P and/or Q493R**.

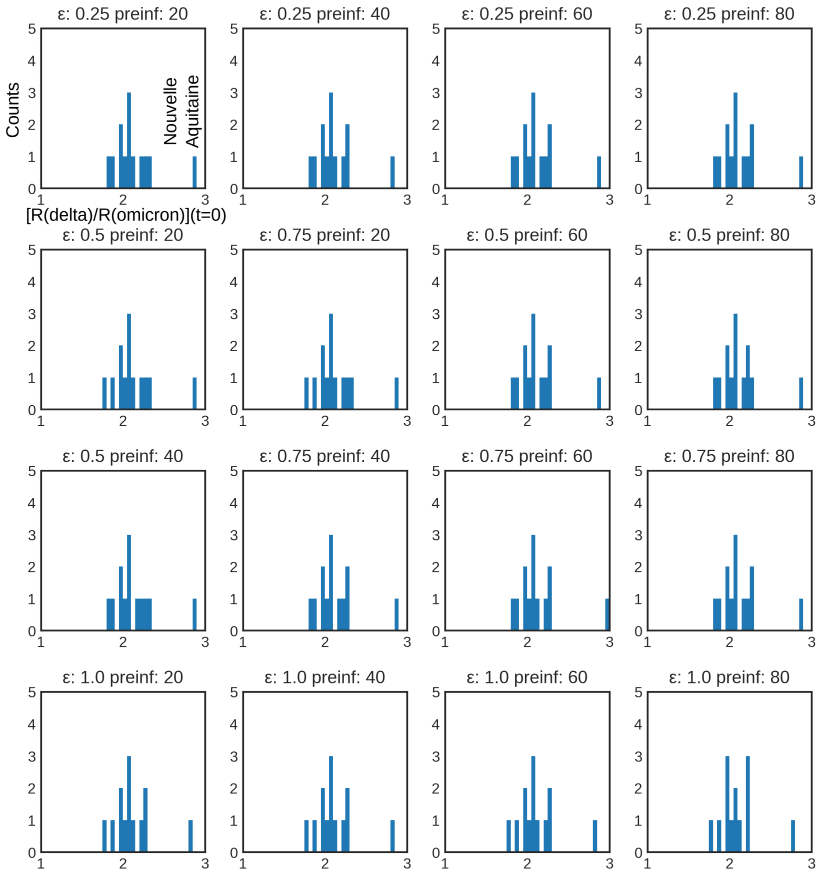


Figure S5: Fitness distribution across regions assuming an omicron generation time of 4 days on average. The distribution of $\frac{\hat{R}_{\text{omicron}}}{\hat{R}_{\text{delta}}}$ by regions shown by a 40 bins histogram between the values 1 and 3 according to generation times of 5 days for the delta variant and 4 days for the omicron variant. The alternative definition of omicron is based on the following mutations: **deletion of site 69/70 and/or substitution K417N and/or S371L-S373P and/or Q493R**.

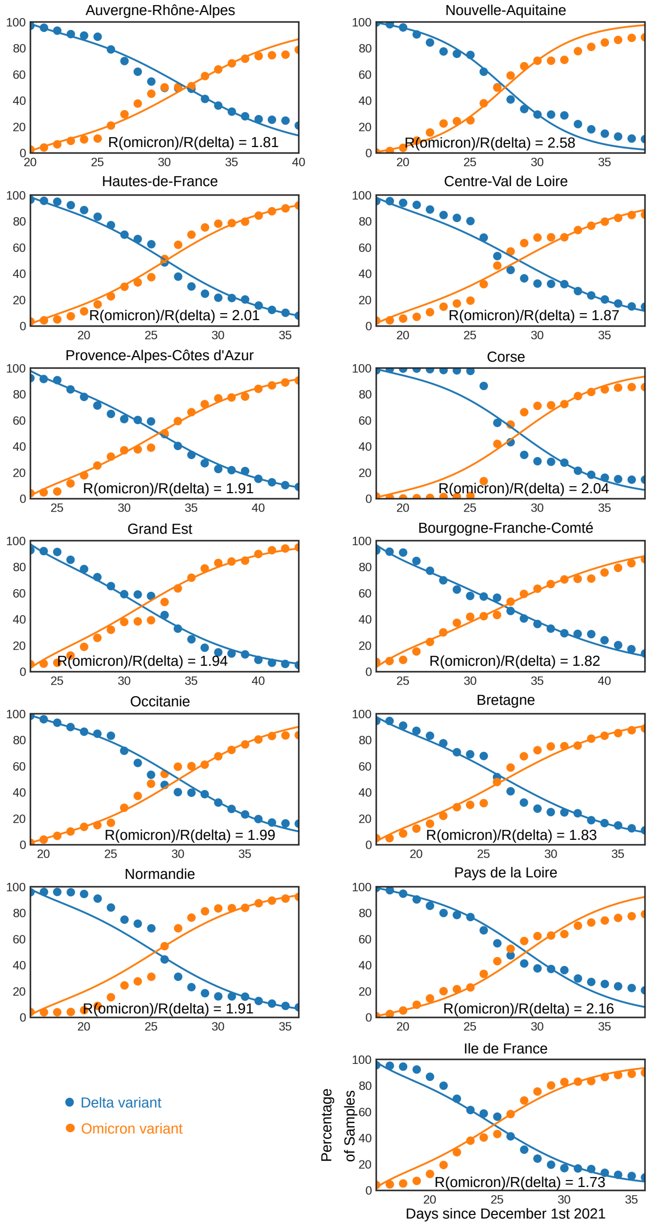


Figure S6: Omicron and delta SARS-CoV-2 variant proportions among positive samples in the regions respective window of opportunity, 10 days before and after the omicron variant exceeds 50%. Dots are representing proportions reported from sampled data published by *Santé Publique France*. Lines represent simulated data with estimated parameter values, here with a delta-omicron cross-immunity of 25% and an initial population that is 80% immunized against the delta variant. The generation time assumed here is 5 days for the delta variant and 3.5 days for the omicron variant. The alternative definition of omicron is based on the following mutations: **deletion of site 69/70 and/or substitution K417N and/or S371L-S373P and/or Q493R**.

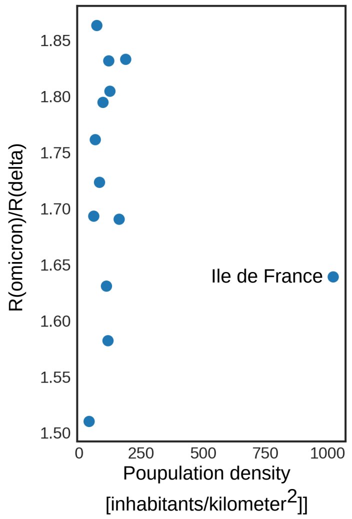


Figure S7: Correlation between population density and estimated relative fitness $\frac{\hat{R}_{\text{omicron}}}{\hat{R}_{\text{delta}}}$ in the different metropolitan French regions. Results shown here were obtained assuming a cross immunity $\epsilon_{\text{omicron}-\text{delta}} = 25\%$ and $a = 80\%$ and a generation time of 3.5 days for the omicron variant. Other parameter choices led to similar results. The definition of omicron is based on the **absence of L452R mutation**.
